# Protocol for a randomised controlled feasibility trial of HD-DRUM, a rhythmic movement training application for cognitive and motor symptoms in people with Huntington’s disease

**DOI:** 10.1101/2023.11.15.23298581

**Authors:** Vasileios Ioakeimidis, Monica E. Busse, Cheney J. G. Drew, Philip Pallmann, Derek K. Jones, Anne E. Rosser, Marco Palombo, Robin Schubert, Claudia Metzler-Baddeley

## Abstract

**Introduction:** Huntington’s disease (HD) is an inherited neurodegenerative disease causing progressive cognitive and motor decline, largely due to basal ganglia (BG) atrophy. Rhythmic training offers promise as therapy to counteract BG-regulated deficits. We have developed HD-DRUM, a tablet-based app to enhance movement synchronization skills and improve cognitive and motor abilities in people with HD. This paper outlines a randomised controlled trial protocol to determine the feasibility of a larger effectiveness trial for HD-DRUM. Additionally, the trial investigates cognitive and motor function measures, along with brain microstructure, aiming to advance our understanding of the neural mechanisms underlying training effects.

**Methods, Design & Analysis:** Fifty individuals with HD, confirmed by genetic testing, and a Total Functional Capacity (TFC) score of 9-13, will be recruited into a two-arm randomised controlled feasibility trial. Consenting individuals with HD will be randomised to the intervention group, which entails eight weeks of at-home usage of HD-DRUM, or a usual-activity control group. All participants will undergo cognitive and motor assessments, alongside ultra-strong gradient (300mT/m) brain microstructural magnetic resonance imaging (MRI) before and after the eight-week period. The feasibility assessment will encompass recruitment, retention, adherence, and acceptability of HD-DRUM following pre-specified criteria. The study will also evaluate variations in cognitive and motor performance and brain microstructure changes resulting from the intervention to determine effect size estimates for future sample size calculations.

**Ethics & Dissemination:** The study has received favourable ethical opinion from the Wales Research Ethics Committee 2 (REC reference: 22/WA/0147) and is sponsored by Cardiff University (SPON1895-22) (Research Integrity, Governance and Ethics Team, Research & Innovation Services, Cardiff University, 2^nd^ Floor, Lakeside Building, University Hospital of Wales, Cardiff, CF14 4XW). Findings will be disseminated to researchers and clinicians in peer-reviewed publications and conference presentations, and to participants, carers and the general public via newsletters and public engagement activities. Data will be shared with the research community via the Enroll-HD platform.

Trial registration & data set: ISRCTN 11906973 (https://doi.org/10.1186/ISRCTN11906973)

Protocol: 01/11/2023 version 1.7

**Strengths and limitations:**

- HD-DRUM is a remotely accessible, tablet-based training tool that can be used at home and allows the objective assessment of adherence and training effects.
- The use of gamification to match users’ practice to a level appropriate to their abilities is expected to increase adherence and acceptability of HD-DRUM by avoiding frustration and boredom due to over- and underchallenge.
- This trial will assess the feasibility of a future fully powered effectiveness randomised controlled trial and any modifications that may need to be implemented in HD- DRUM.
- The randomised controlled study design will allow the estimation of training-induced variability of changes in clinical and brain imaging measures, using state-of-the art ultra-strong gradient 3T MRI.
- Due to the nature of the intervention and limited resources, researchers and participants are not blinded to group allocation.

## INTRODUCTION

### Background and Rationale

Huntington’s disease (HD) is a genetic neurodegenerative disease that leads to the progressive loss of cognitive and motor abilities, largely due to basal ganglia (BG) atrophy. Striatal atrophy^1^ and microstructural alterations in white matter (WM) connections^2^ with associated cognitive control dysfunctions^3–5^ occur many years before the clinical onset of motor symptoms and will eventually negatively impact a person’s ability to live independently^6^.

Currently, there are no disease-modifying treatments for HD and symptomatic therapies are limited with significant side effects^7^ ^8^. There are no HD-specific cognitive interventions and only very few studies have investigated cognition-oriented interventions in people with HD^9^.

Rhythmic auditory stimulation (RAS) is a form of neurologic music therapy (NMT)^10^ that uses rhythmic beats as external cues to trigger movements^11^. RAS has been found to improve gait and mobility in people with Parkinson’s disease (PD)^11–13^ and holds potential as an intervention for improving attention and executive functions^12^ ^14^. While PD and HD are clinically distinct diseases, they are both associated with BG neurodegeneration and deficits in interval timing and spontaneous rhythm generation^15^. The BG are considered to play a key role in the prediction of upcoming events by internally generating temporal pacing signals^16–18^. Together with the cerebellum and cortical areas involved in sensory, motor, and attention processing, they form an extended network of brain regions that supports rhythmic sensory processing and movement generation^16^. RAS interventions are proposed to work by compensating for the loss of BG-generated timing and rhythm signals with external rhythmic cueing^11^. However, the actual neural mechanisms of rhythmic processing as well as the clinical effects of RAS interventions in people with HD remain unknown.

Previously, we explored eight weeks of ∼15 minutes of bongo drumming five times per week as a therapeutic tool for people with HD^19^ ^20^. We observed performance improvements in executive function tasks and changes in WM microstructural measurements (including an estimate of myelin) of callosal and cortico-putamen connections^19^ ^20^. Although these initial pilot results are promising, they are preliminary in nature and necessitate validation through a fully powered randomised controlled trial (RCT). This study aims to investigate the feasibility of a larger RCT, as well as to explore any indicative effects and the neural mechanisms that may be responsible for potential clinical advantages associated with RAS.

For this purpose, we have co-designed with people with HD, a tablet-based RAS training application (app) (HD-DRUM)^21^ that is suitable for clinical evaluation because it allows the accurate quantification of training progression and adherence and uses gamification to match individual performance levels to training difficulty with the aim to increase acceptability. As HD-DRUM can be used at home it holds the potential of expanding intervention delivery to greater participant numbers for an effectiveness trial.

Here we describe the protocol of an RCT to determine the feasibility of a larger effectiveness RCT into the clinical effects of HD-DRUM in people with HD. The trial will also explore training-induced changes in cognitive and motor functions as well as in magnetic resonance imaging (MRI) measures of morphology and microstructure of cortical and subcortical brain regions involved in rhythmic processing (BG, cerebellum, auditory and motor cortices). The clinical and neural effects of eight weeks of HD-DRUM will be compared with a usual-activity control group in people with HD that will allow us to explore whether HD-DRUM can improve on current practice.

HD-DRUM has the potential to be a future training tool for slowing cognitive and motor decline in people with HD, alongside candidate disease-modifying treatments for HD that are currently under consideration. Even a small delay of disease progression would translate into direct benefits for the quality of life of patients and their families.

### Primary objective

To assess the feasibility of a larger effectiveness RCT investigating eight-weeks of at-home HD-DRUM intervention compared with usual activities in people with HD.

### Secondary objectives

To gain estimates of variability and absolute change before and after eight weeks of HD-DRUM compared to usual-activity control in the following measures:

1. Performance measures in cognitive and motor tasks for sample size calculations for a future RCT.
2. The means of grey and white matter microstructure in BG and cerebellar brain networks^3^ to explore the neural mechanisms that underpin any training effects.
3. The means of performance measures in cognitive and motor tasks and of grey and white matter microstructure in BG and cerebellar brain networks^3^ to inform about disease-specific neural mechanisms including compensatory processes.

### Trial design

We will conduct a two-arm randomised controlled feasibility trial of individuals with HD randomised to eight weeks of HD-DRUM intervention or usual-activity control (Figure 1).

**Figure 1:**
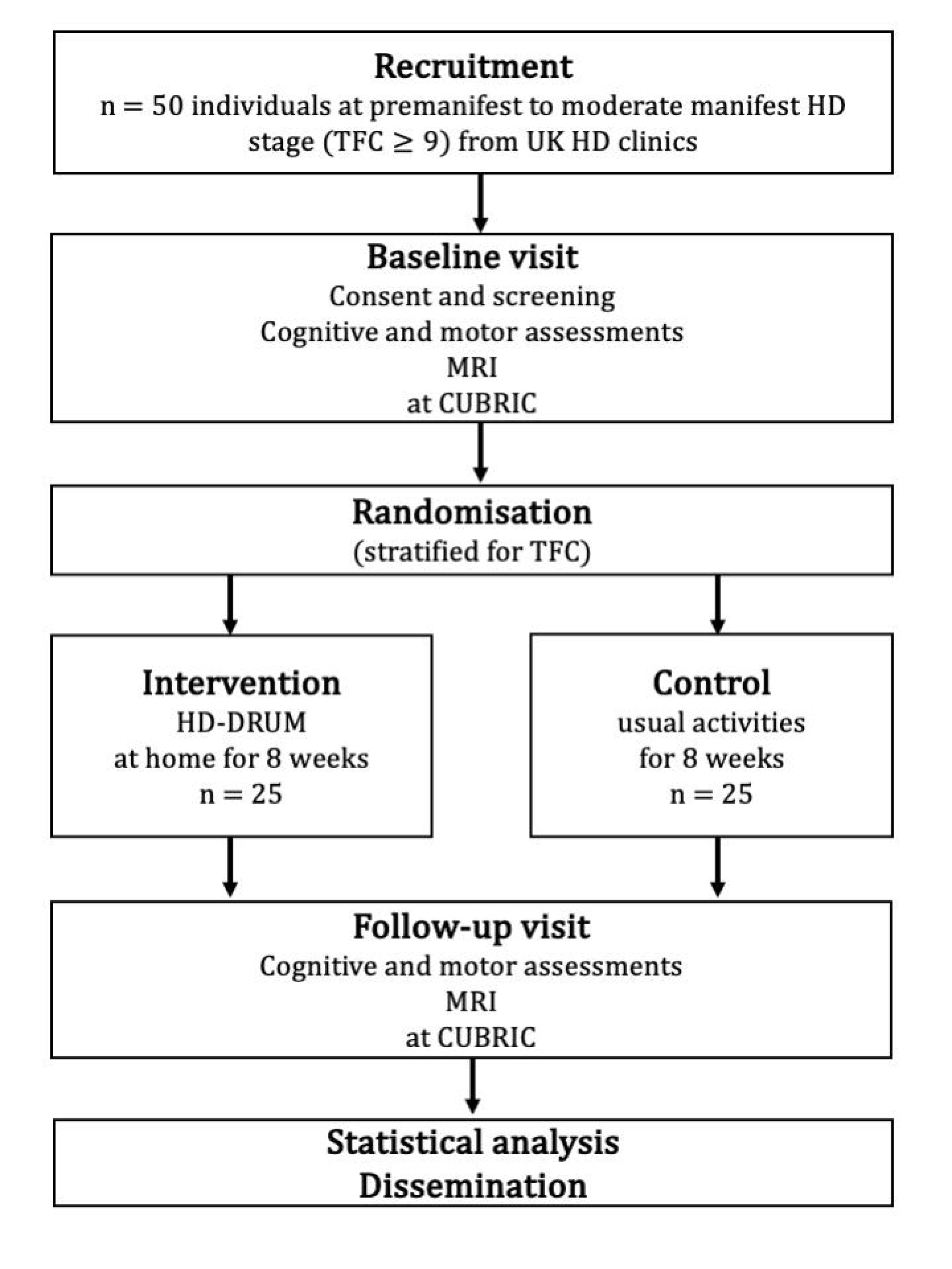
Schematic overview of the two-arm randomised controlled feasibility trial.

## METHODS

### Participants, interventions, and outcomes

#### Study setting

Participants will be identified, approached, and recruited via specialist HD clinics in the UK. All baseline and follow-up clinical and MRI assessments will take place at the Cardiff University Brain Research Imaging Centre (CUBRIC). Participants will engage individually with the HD-DRUM intervention at home.

#### Recruitment

Eligible individuals will be identified by their clinician during their annual Enroll-HD visit. Individuals interested in the trial will receive the study information sheet. They will be asked to provide consent for the researchers to contact them to find out if they wish to proceed with the study. If they decide to participate, during their first appointment at CUBRIC they will be asked to provide informed written consent and complete the study assessments. Participants will be reimbursed for their travel expenses to CUBRIC as well as overnight accommodation in Cardiff if they wish to.

The study will also be advertised on posters in waiting rooms of participating clinics and via newsletters and websites of Health and Care Research Wales and the Huntington’s Disease Association UK. In these instances, interested parties must first contact one of the participating HD centre teams for further information.

#### Eligibility criteria

Participants will be eligible to take part in the study if they meet all of the inclusion criteria and none of the exclusion criteria as specified below.

#### Inclusion criteria

Individuals over the age of 18 years with a good command of the English language who fulfil the following criteria:

1 premanifest or manifest HD as confirmed by genetic testing for the presence of the mutant huntingtin allele,

2 a Unified Huntington’s Disease Rating Scale (UHDRS) Total Functional Capacity (TFC)^22^ score between 9 and 13,

3 who participate in the Enroll-HD^23^ observational study (former Registry) (REC no 04/WSE05/89) or in routine clinical assessment of their TFC score.

#### Exclusion criteria

A history of any other neurological condition and/or an inability to provide informed consent. MRI contra-indications (e.g., pacemakers, stents) will preclude participation in the MRI but not the training part.

#### Intervention

The HD-DRUM app is a computerised version of the drumming training intervention we have previously devised^19^ ^20^. A detailed description of HD-DRUM according to the template for intervention description and replication (TIDieR)^24^ has been published^21^.

In brief, the app consists of twenty-two ∼10-15 minutes audio training sessions (Figure 2A) that introduce rhythmic patterns of different styles including paradiddles, hip-hop, funk, and reggaeton with and without a background metronome or music. Participants will be encouraged to drum along with the audio instructions on two virtual drums (Figure 2B) on the tablet screen; with their left hand on a blue triangle and with their right hand on a red circle. The virtual drums produce visual (shrinking) and audio feedback (high and low pitch bongo sounds) when tapped. The patterns gradually increase in complexity and tempo through the programme. Initially, participants will practise with each hand separately before drumming with both hands, starting with one hand first and then reversing the order of the hands. Participants will be asked to complete a training session a day, five times a week, for eight weeks (40 session in total). They will be able to move through the programme at their own pace and repeat each session as often as they wish. Response time and accuracy will be measured within the app and a success threshold of 70% will need to be achieved to move on to the next level. Researchers will remotely assess training adherence by monitoring coded output files generated and uploaded to a project-specific space on Google Firebase (Google LLC) (https://firebase.google.com) when the tablet is connected to the internet.

**Figure 2:**
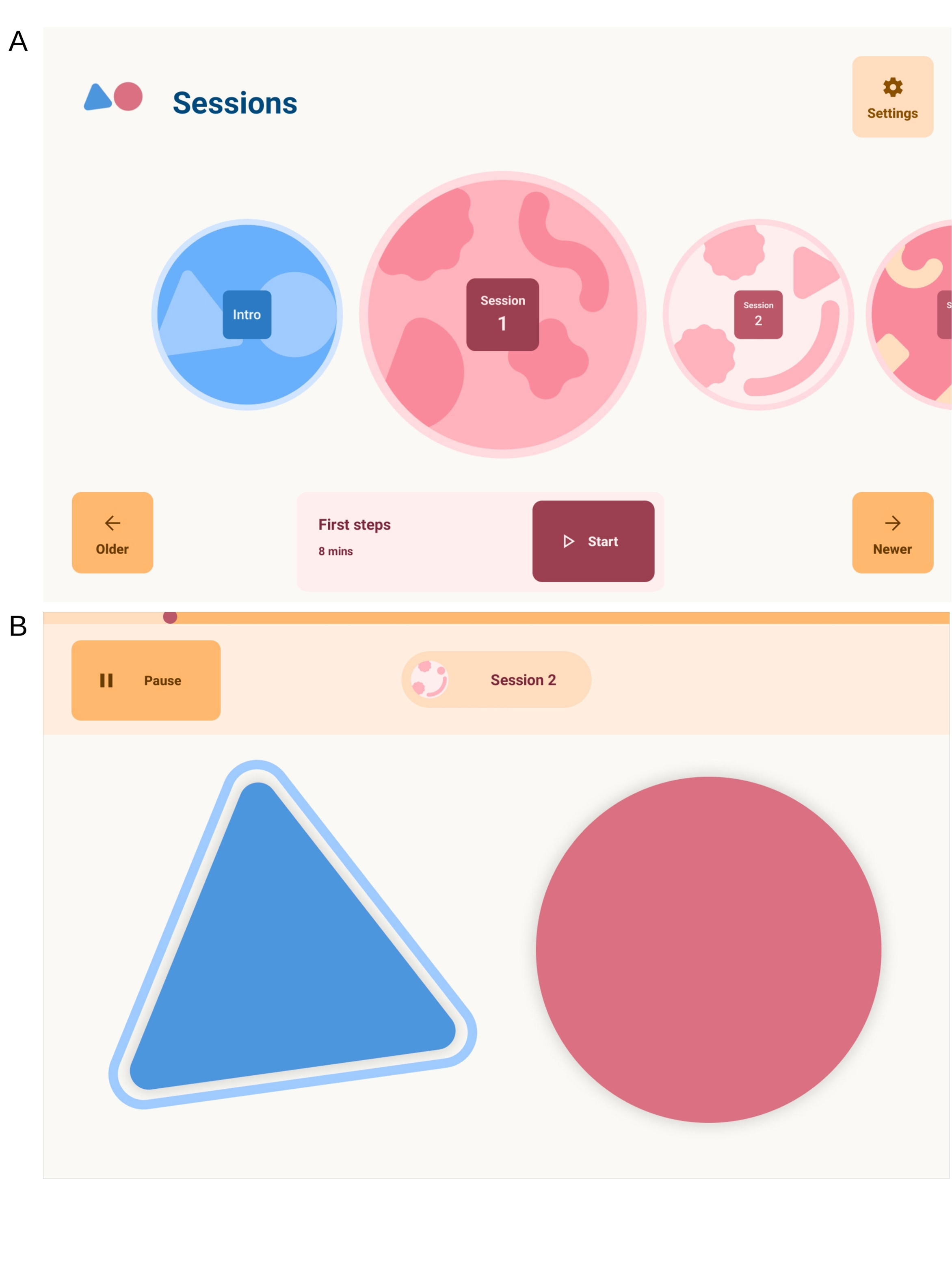
The HD-DRUM app environment. Panel A) shows the app home screen interface that allows users to navigate through and select unlocked training sessions. Panel B) shows the virtual drums, a blue triangle (high pitch bongo) and red circle (low pitch bongo), participants use to tap in synchronization with the audio instructions.

Participants in the intervention group will be provided with a tablet with the HD-DRUM app installed and an instruction manual for home use. Following completion of baseline assessments participants will receive a face-to-face introduction to the app to familiarise themselves with HD-DRUM. To support retention to the intervention, researchers will maintain weekly contact with participants to provide feedback, assist with any queries or problems, and help remove potential barriers. Participants in the control condition will be offered the opportunity to borrow a tablet with HD-DRUM to try the training out after they have completed the final assessment of the study.

### Outcome assessments

#### Primary feasibility outcome assessments

1. Recruitment will be measured as the number of participants assessed for eligibility, eligible, approached, and consented into the study. Reasons for not approaching or enrolling eligible participants will be recorded on a screening log.
2. Retention will be measured by the number of participants who complete the study after attending the follow-up visit after eight weeks. Reasons for withdrawal and loss to follow-up will be recorded where possible.
3. Acceptability will be measured with a semi-quantitative self-report questionnaire on rating how engaging the training was, appropriateness of frequency, duration, and difficulty, perceived beneficial or detrimental training effects, as well as likes, dislikes, and potential improvements within the app. This will inform about any further app adjustments required to address the needs of people with HD for a future RCT.
4. Adherence to the training will be automatically tracked within HD-DRUM by recording the frequency and duration of training sessions.

Feasibility (recruitment, retention, adherence) success criteria have been predefined according to the following traffic lights system: Green (all feasibility rates ≥ 70%) will signal that the trial was successful, amber (no rates below 40% but at least one below 70%) that the project requires review and design changes, and red (at least one rate < 40%) that a future RCT will not be feasible.

#### Secondary outcome measures

Performance in the following measures will be assessed before and after eight weeks of intervention or usual-activity control (see Table 1 for participant timeline).

**Table 1.** Schematic diagram of participant timeline.

| Assessments for all participants |  |  |  |
| --- | --- | --- | --- |
|  | Baseline study visit | HD-DRUM training intervention (completed at home with support and reminders) or control time period | Follow-up study visit |
|  | Day 1 | 8-week intervention period | End of week 8 (+ 2 weeks) |
| Cohort Descriptive information |  |  |  |
| MOCA<sup>†</sup> | X |  |  |
| TOPF | X |  |  |
| Self-report questionnaire |  |  |  |
| Patient Health Questionnaire-8 | X |  | X |
| HD-DRUM participant evaluation survey |  |  | X |
| Paper and pencil tasks |  |  |  |
| Stroop task | X |  | X |
| Verbal fluency | X |  | X |
| SDMT | X |  | X |
| Categorical fluency | X |  | X |
| Verbal trails (letter-digit switching) | X |  | X |
| Computerised tasks |  |  |  |
| N-back Audio (letters) | X |  | X |
| N-back V isuospatial (square changing positions in grid) | X |  | X |
| N-back dual (audio and visuospatial) | X |  | X |
| Flanker task | X |  | X |
| Q-Motor assessments |  |  |  |
| Speeded finger tapping (left/right index finger) | X |  | X |
| Finger metronome tapping (left/right index finger) | X |  | X |
| Pointing and tapping (dominant hand) | X |  | X |
| Dual (pointing and tapping with dominant hand, speeded finger tapping with opposite hand) | X |  | X |
| Speeded foot tapping (left/right) | X |  | X |
| Metronome foot tapping (left/right) | X |  | X |
| Brain imaging |  |  |  |
| T1-weighted MPRAGE | X |  | X |
| msHARDI | X |  | X |
| qMT | X |  | X |
| mcDESPOT | X |  | X |
| HD-DRUM group |  |  |  |
| HD-DRUM training intervention supported by email and telephone reminders |  | X |  |
| Control group |  |  |  |
| Standard care, usual activities |  | X |  |

Abbreviations: **HD**: Huntington's disease; **mcDESPOT**: Multicomponent Driven Equilibrium Single Pulse Observation of T1 and T2; **MOCA**: Montreal Cognitive Assessment; **msHARDI**: Multi-shell High Angular Resolution Diffusion Imaging; **qMT**: Quantitative Magnetization Transfer; **SDMT**: Symbol Digit Modalities Test; **T1-MPRAGE**: T1-weighted Magnetisation Prepared-Rapid Gradient Echo Sequence; **TOPF**: Test of Premorbid Functioning.

#### Cognition, motor function, and mood

Participants will be assessed in the following tests that have been shown sensitive to cognitive^6^ and motor symptoms in HD^25^ ^26^. The testing session will take approximately two hours including breaks.

1. Fine motor functions of speeded finger and foot tapping, tapping in synchronization with a metronome and continuation without as well as dual-task motor ability of simultaneously tapping with one hand and pointing with the other will be assessed with the Quantitative Motor (Q-Motor) assessment test battery^25^. Mean and standard deviation (SD) of tapping and pointing speed will be measured in seconds, mean and SD tapping force will be measured in Newtons.
2. Cognitive functions, notably executive abilities of attention switching, distractor suppression, and working memory updating^27^ will be assessed with paper-and-pencil and computerised tasks from the Psychology Experiment Building Language (PEBL) test battery^28^ and implemented in MATLAB ^29^ ^30^. Table 2 provides details about all cognitive tasks including their outcome measures and cognitive domain of interest.
3. Mood will be assessed with the brief self-report Patient Health Questionnaire (PHQ-8)^31^.

**Table 2.**
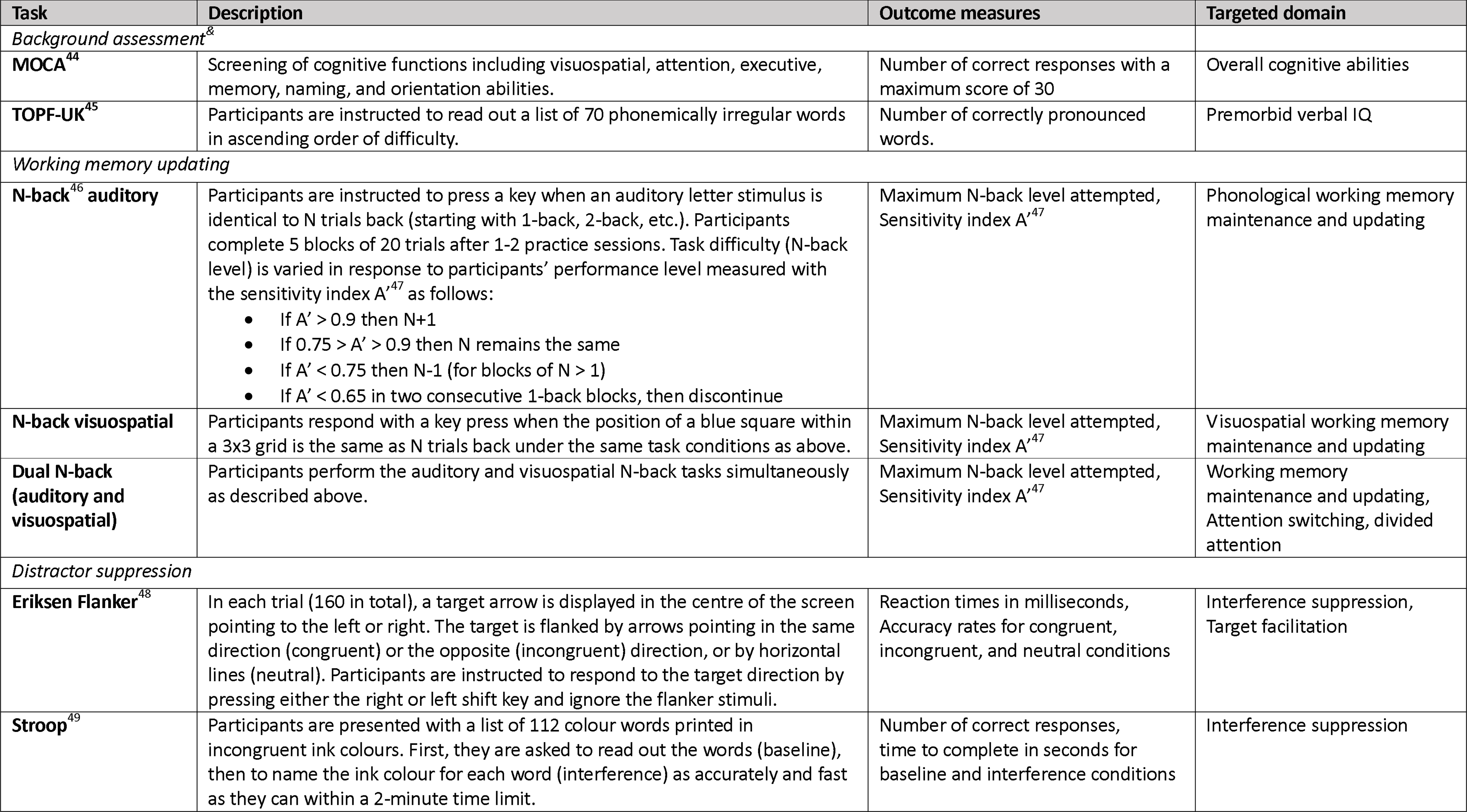
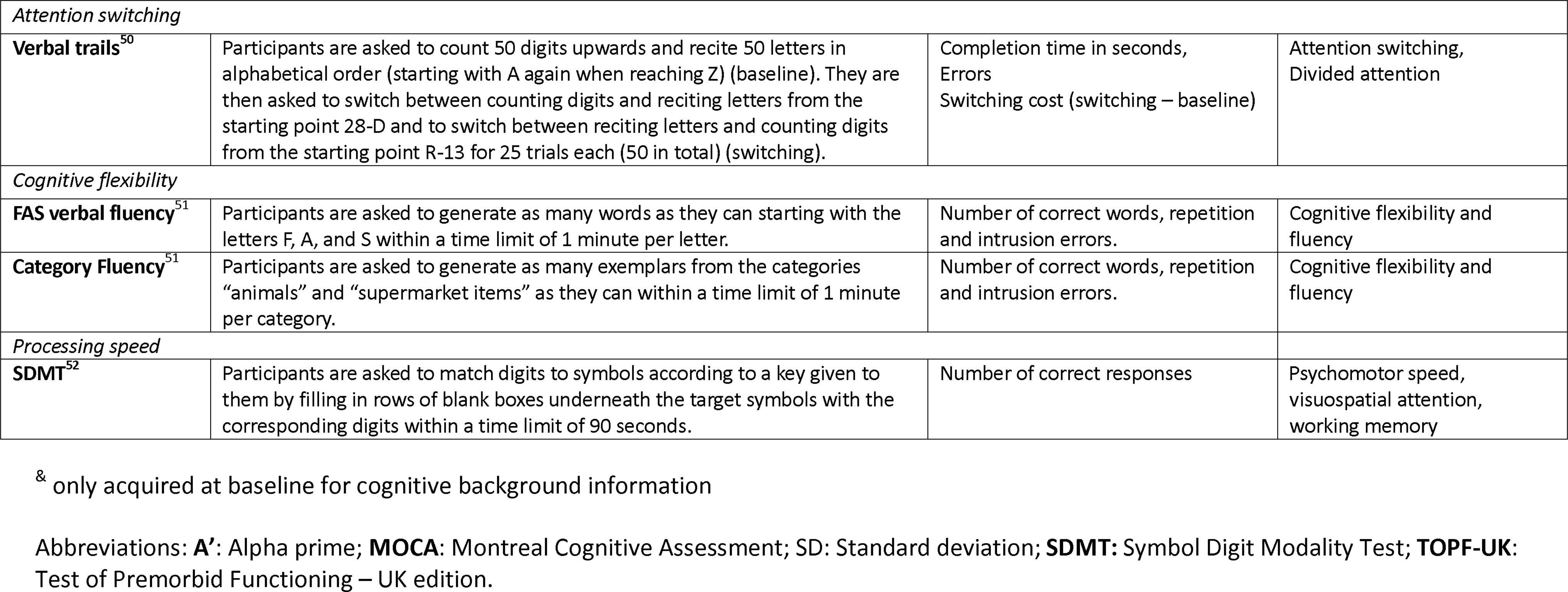
Overview of cognitive assessments.

#### MRI brain morphology and microstructure

MRI data will be collected on the UK National Microstructural Imaging Facility, a 3 Tesla MRI Siemens Connectom system with ultra-strong (300mT/m) gradients. Ultra-strong gradients allow the acquisition of diffusion-weighted data with high diffusion-weighting b-values, with good signal to noise ratio for the estimation of intracellular diffusion properties^32^. Multi-shell high angular resolution diffusion imaging (msHARDI)^33^ data (max b-value = 6,000 s/mm^2^) will be acquired to estimate microstructural white and grey matter properties of axon and soma density^34^ ^35^. Myelin properties will be estimated with quantitative magnetization transfer (qMT) imaging^36^ and multicomponent driven equilibrium single pulse observation of T and T (mcDESPOT)^37^. Table 3 provides details of the MRI protocols including their acquisition parameters, the outcome index maps from the analysis based on biophysical modelling, and estimated white and grey matter tissue properties.

**Table 3.**
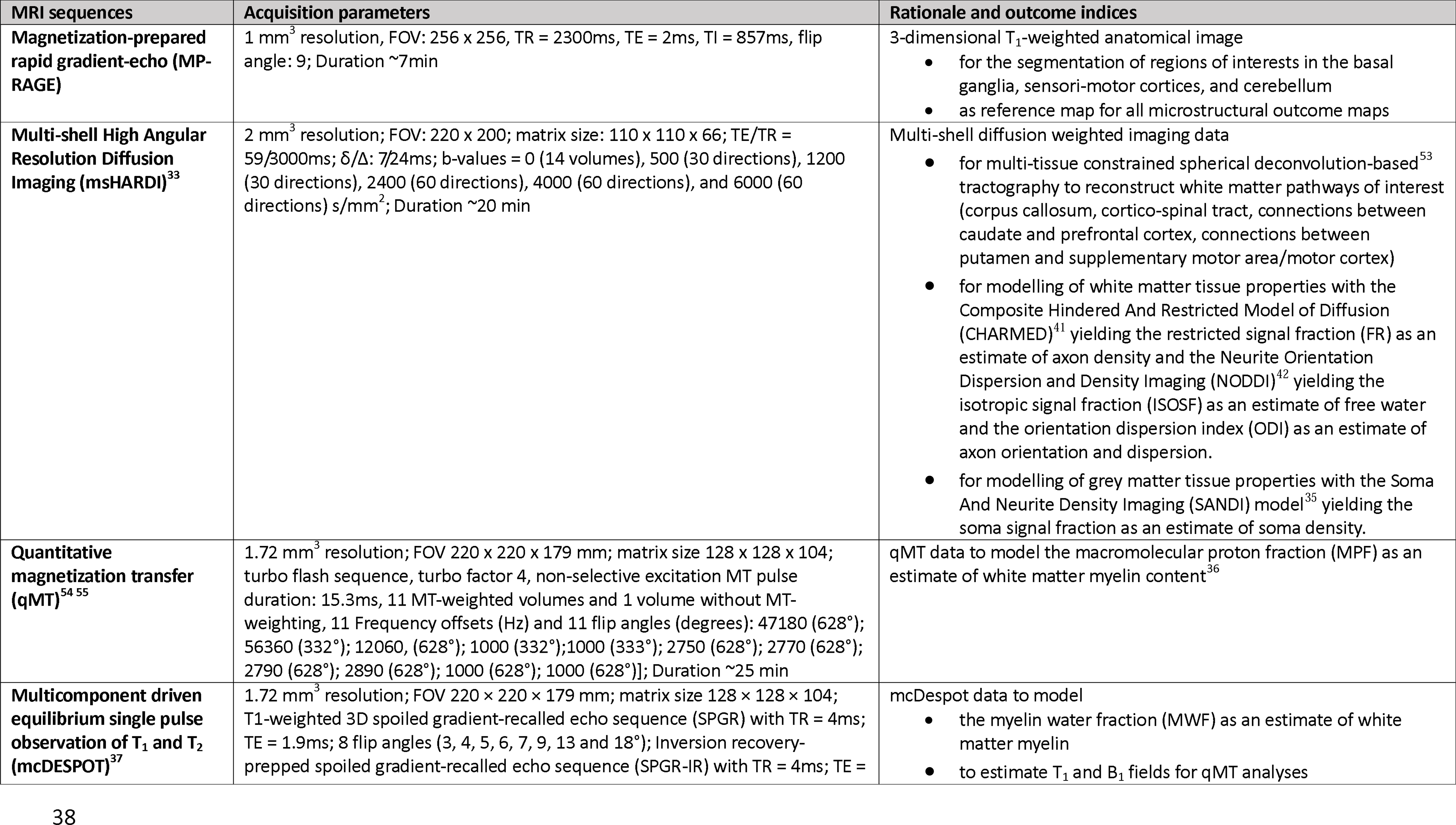
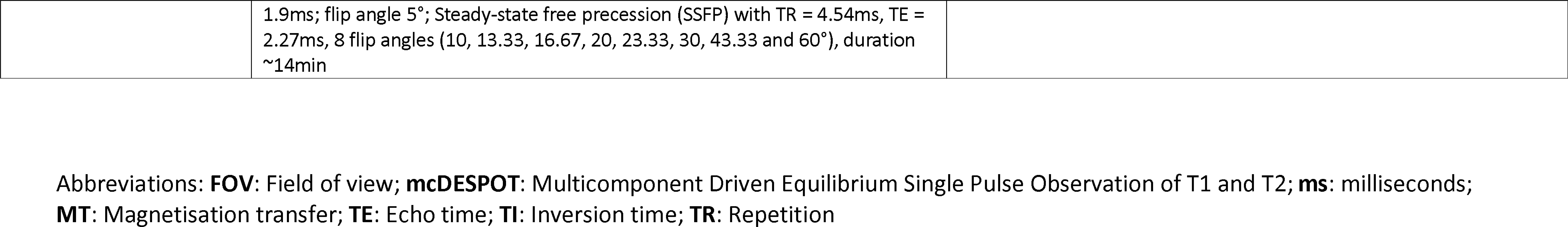
Overview of Magnetic Resonance Imaging data.

All MRI protocols have been piloted in individuals with HD. They take 60 minutes in total to acquire and are well-tolerated. Our previous research has identified microstructural differences in HD patients compared with healthy controls^38^ ^39^. Importantly, we found drumming training-induced increases in the qMT-based macromolecular proton fraction (MPF) in callosal white matter suggestive of increased myelin^19^.

#### Sample size

Following the Consolidated Standards of Reporting Trials (CONSORT) statement extension to randomised pilot and feasibility trials^40^, no formal power calculations have been conducted. We aim to recruit 50 individuals with HD. This number is pragmatic given the size of the target population, level of engagement required, and resources available. It will allow estimation of recruitment, retention, and adherence rates with a 95% binomial confidence interval of no more than plus or minus 15 percentage points irrespective of the point estimate.

The combined population of the participating patient identification centres should be adequate to achieve recruitment target, but this assumption will be tested as part of the feasibility assessment. While there is facility to add more patient identification centres, further strategies to enhance recruitment will not be used within this feasibility trial.

### Assignment of interventions

#### Allocation

Randomisation will be stratified for TFC score to ensure comparability of the groups with regards to disease burden. There will be seven strata corresponding to TFC scores of 9, 10, 11, 12 and 13, respectively. One randomised group allocation sequence per stratum will be computer generated based on pseudo-random numbers by a statistician (PP) independent to patient recruitment and data collection, using R version 4.1.0^41^ and uploaded onto the HD-DRUM project specific Research Electronic Data Capture (REDCap)^42^ site.

#### Blinding (masking)

The researchers conducting the baseline assessments (cognitive, motor, and MRI) have no access to the randomisation list implemented in REDCap and will be blind to the group allocation until baseline assessments are completed. However, once group allocation has taken place at the end of the baseline assessments, blinding of the researchers involved in the data collection and participants will not be possible due to the nature of the intervention and control conditions and due to limited resources and staffing. The analysis of the Q-Motor data will be conducted blind to group allocation by an independent researcher not involved in the data collection (RS). The lack of blinding of the clinical and MRI outcomes is a limitation of the study design, but quantitative outcome measures have been used where possible to minimise any potential biases that may be introduced due to a lack of blinding.

### Data collection, management, and analysis

#### Data collection methods

The project will be conducted following Good Clinical Practice, data protection in accordance with the Data Protection Act 2018, and CUBRIC standard operating procedures (SOPs) including MRI safety and operation guidelines. The project will adhere to any applicable Centre for Trials Research (CTR) SOPs, including those for project management, data management and protection, and adverse/serious adverse event reporting. Research staff (VI) will receive training in the above.

Training-specific performance improvements, namely speed and accuracy of drumming will be recorded by the HD-DRUM app. Frequency and duration of training engagement will also be recorded by HD-DRUM as a measure of adherence.

Cognitive and motor performance measures will be collected on paper data collection forms and electronically via PEBL and MATLAB on a personal computer in a quiet testing room in CUBRIC. Electronic data capture of Q motor data is automated by the Q-Motor data acquisition software Q-MedX.

MRI data will be acquired on the 3 Tesla MRI Siemens Connectom system at CUBRIC.

#### Data management

Participants will be assigned a study and CUBRIC ID and all electronic and hard-copy data will be coded with these IDs. Electronic data will be stored within Cardiff University’s firewall and password-protected computer systems. Cognitive and motor clinical outcome and training data (stored in .csv files) will be managed with the REDCap web application (www.project-redcap.org) that allows secure storage, management, and sharing of data using a confidential security level. Paper record forms will be scanned, and electronic copies will be uploaded onto RedCAP. Cognitive and motor scores are entered by the researcher (VI) and will be quality checked against the original record forms by other members of the research team.

The data collection with the drumming training application complies with General Data Protection Regulation (GDPR) and data privacy regulations. No personal or identifiable user information is stored on the tablet or on Google Firebase. The drumming training output files are coded with a sixteen-digit-letter-string tablet ID. Firebase services encrypt data at rest and during transit using hypertext transfer protocol secure (HTTPS).

MRI data will be stored securely on the CUBRIC partition of XNAT (www.central.xnat.org). Study and CUBRIC ID keys and any personal information will be stored for the duration of the project securely in a locked cupboard in the Principal Investigator’s (CMB) office in CUBRIC. Only researchers associated with the study will have confidential access to files, which allow the matching of recorded data to participants. No data, whether paper or electronic, will leave Cardiff University sites without being completely coded, i.e., all identifiable data will be removed from the dataset. Any electronic files or disks will be stored on Cardiff University sites and electronically on Cardiff University systems. At the conclusion of the study participant identifiable data will be destroyed and non-identifiable data will be archived, although it will still be accessible to the study team. Data will be archived for 15 years, in line with Cardiff University policies and procedures.

#### Statistical methods

The study will be reported in accordance with the CONSORT statement extension to randomised pilot and feasibility trials^40^. As this is a feasibility trial, it is not formally powered to test for effectiveness of the intervention whilst controlling type I error. The primary aim is that of assessing feasibility, recruitment, retention and adherence rates and acceptability rating scores and thematic responses will be documented and evaluated according to the success criteria described above. Feasibility percentage rates will be calculated as follows:

- Recruitment rate = 100 * (number of participants consented / number of participants eligible and approached) %
- Retention rate = 100 * (number of participants completing the study at eight-week follow-up/ number of participants consented) %
- Adherence rate (frequency) = 100 * (number of days app used for training / 40 days) %
- Adherence rate (duration) = 100 * (number of minutes app used for training /(40 days * 11.3 minutes average session duration = 451 minutes) %

Descriptive summaries such as mean and SD estimates (for continuous variables) of effect sizes and 95% confidence intervals will be calculated for all secondary clinical and MRI outcome measures. Motor and mood measurements are detailed in the outcome section above and information about the cognitive and MRI outcome measures can be found in Tables 2 and 3. Differences in mean and SD of participants’ absolute and percentage changes from baseline will be calculated to provide estimates of effect size and variability of any HD-DRUM-induced changes between individuals with HD in the HD-DRUM and those in the usual-activity control group. All calculations will be conducted in SPSS^43^ and R^41^. This will provide information to help estimate the sample size required for a future definitive RCT into HD-DRUM in this patient population. Note that the absence of a change in individuals with HD in the HD-DRUM group compared with the HD control group may suggest a beneficial effect of the intervention as HD is a progressive neurodegenerative disease.

### Monitoring

#### Data monitoring and ethics oversight

This feasibility trial is expected to pose minimal risk to participants, and hence no formal Data Monitoring Committee has been established. However, the research team will produce regular data monitoring reports for the Project Steering group, comprised of a trial statistician, clinician, and public involvement member, who are independent of the project management team, and will assess and monitor the data quality on an annual basis throughout the study.

#### Harms

MRI scanning is non-invasive and has no known significant adverse health effects when appropriate screening and safety measures are in place and implemented. CUBRIC SOPs will be followed to ensure this. Any adverse effects including rare events of peripheral nerve stimulation, dizziness, mild nausea, a metallic taste in the mouth, and the sensation of seeing flashing lights during the MRI scanning will be monitored, recorded, and documented. These side effects, if experienced, may be uncomfortable but resolve after leaving the magnetic field and participants will be warned at the beginning of the MRI scanning session. It will also be explained to participants that the researchers do not have expertise in MRI diagnosis, as they are not medical doctors. Participants should not regard the research scans as medical screening procedures and if they had any health concerns, they should contact their medical practitioner. In the unlikely event of an unexpected finding, a neurological consultant (AR) will be asked to examine the scans, and if appropriate to report back to the participant’s GP.

Finally, in the unlikely event of adverse events occurring from the HD-DRUM intervention or the cognitive and motor assessments, these will also be monitored, recorded, and reported.

### ETHICS AND DISSEMINATION

#### Research ethics approval

The study will be conducted in accordance with the recommendations for physicians involved in research on human participants adopted by the 18th World Medical Assembly, Helsinki 1964 and later revisions. Ethical approval for this feasibility study was given by the Wales Research Ethics Committee (REC) 2 (22/WA/0147). Cardiff University has agreed to act as sponsor for the study (SPON1895-22).

All amendments to the ethical approval will be recorded in the trial registration and any future amendments to be made will be done in accordance with the Integrated Research Application System (IRAS) regulation and will be communicated to sites via the UK permissions system.

#### Consent

Informed written consent will be sought by means of participant dated signature and dated signature of the person who obtained the informed consent on the consent form provided in the Appendix. The participant will be allowed as much time as needed to consider the patient information sheet and given the opportunity to ask the researcher or independent parties any questions to decide whether they will participate in the study. All participants will have the procedures explained to them in detail, including that the study design is randomised, and be reminded that they can withdraw at any time, without the need to give a reason. The original signed consent form will be retained at the study site in CUBRIC, and participants will receive a copy.

#### Declaration of interests

The authors have no conflicts of interests to declare.

#### Access to data

This project has been endorsed by the Enroll-HD Scientific Oversight Committee (SOC) (14/11/2022) and the use of Enroll captured data has been approved for this trial. At the end of the project, the coded study data will be shared and made accessible to the research community via the Enroll-HD specific data request process.

#### Dissemination poli3cy

Dissemination activities to share the progress and results of our projects with people affected by HD will be led by our Public Involvement (PI) contributors. Dissemination strategies will include information sessions for people affected by HD, clinicians, and the general public as well as communications via social media, newsletters and websites of the HD Association (HDA) UK, the European Huntington Disease Network (EHDN) and HCRW. The findings of this trial will be disseminated as scientific publications and presentations at conferences including at the biennial EHDN plenary meeting. Authorship eligibility will be assessed following the International Committee of Medical Journal Editors (ICMJE) guidelines.

### Patient and Public Involvement

This study was developed and planned together with members of HD Voice, the PPI group of the UK HD Association comprised of people living with HD, their families, and carers. PPI members are involved in the management and oversight of the study as part of the study management and steering groups. The HD-DRUM intervention was co-designed with people with HD. PPI members will contribute to the interpretation and the dissemination of the study findings.

## Author contribution statement

Conceptualisation: CMB, AR, MB, CD, PP; Original draft preparation: CMB, VI; Review and editing: CMB, VI, AR, MB, CD, PP, DJ, MP, RS; Data curation: CMB, VI; Formal analysis: CMB, VI, PP, MP, RS; Interpretation: CMB,VI, AR, MB, CD, PP, MP, DJ, RS; Funding acquisition: CMB. All authors have read and agreed to the published version of the manuscript.

## Data Availability

Data will be shared with the research community on request via the Enroll-HD platform.

## Acknowledgements

We would like to thank the clinical and administerial staff at the participating patient identification centres for their help with identifying suitable patients for the study. We would like to thank Eileen Donovan, Kim Munnery, and Jane Davies from the Cardiff HD clinic; Natalie Rosewell, Anya Soonderpershad, and Dr Liz Coulthard from the Bristol Brain Centre; Claire Tilley, Jessica Prado Mota, and Dr Timothy Harrower from the Royal Devon University Healthcare NHS Foundation Trust in Exeter; Jennifer De Souza and Dr Hugh Rickards from the Birmingham and Solihull Mental Health NHS Foundation Trust; and Jenni Burns, Dr Sundus Alusi, and Dr Rhys Davies from the Walton Centre NHS Foundation Trust in Liverpool. We would like to thank Lucy Layland and Amy Dangerfield for their help with data collection. In addition, we would like to thank all Public Involvement contributors and the members of the Enroll Scientific Oversight Committee for their input into the study as well as all participants for their generous time commitment to help us assess the feasibility of HD-DRUM.

## Funding

This work was supported by a National Institute for Health Research (NIHR) and Health and Care Research Wales (HCRW) Advanced Fellowship to CM-B (grant number: NIHR-FS(A)-2022).

## Appendix

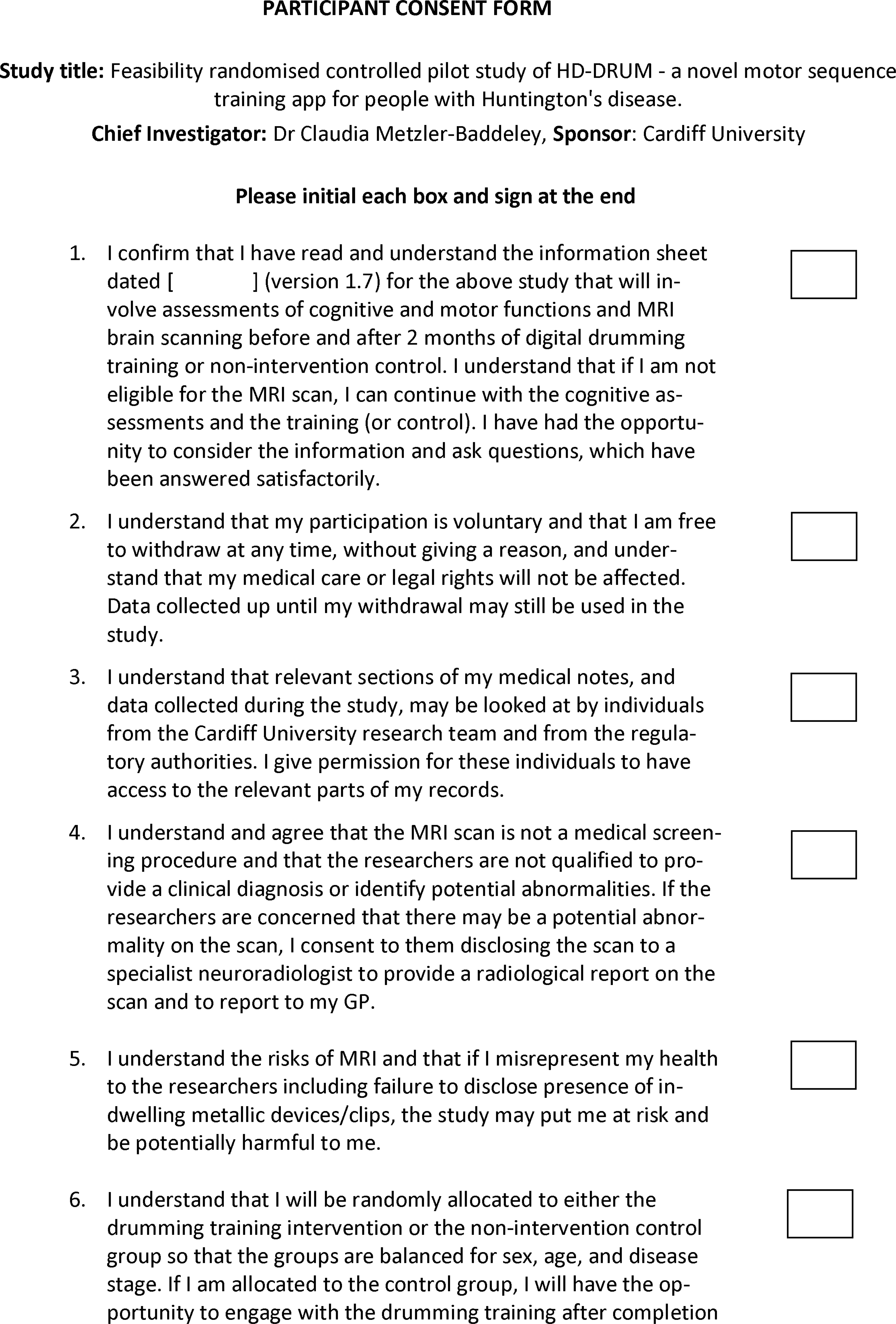
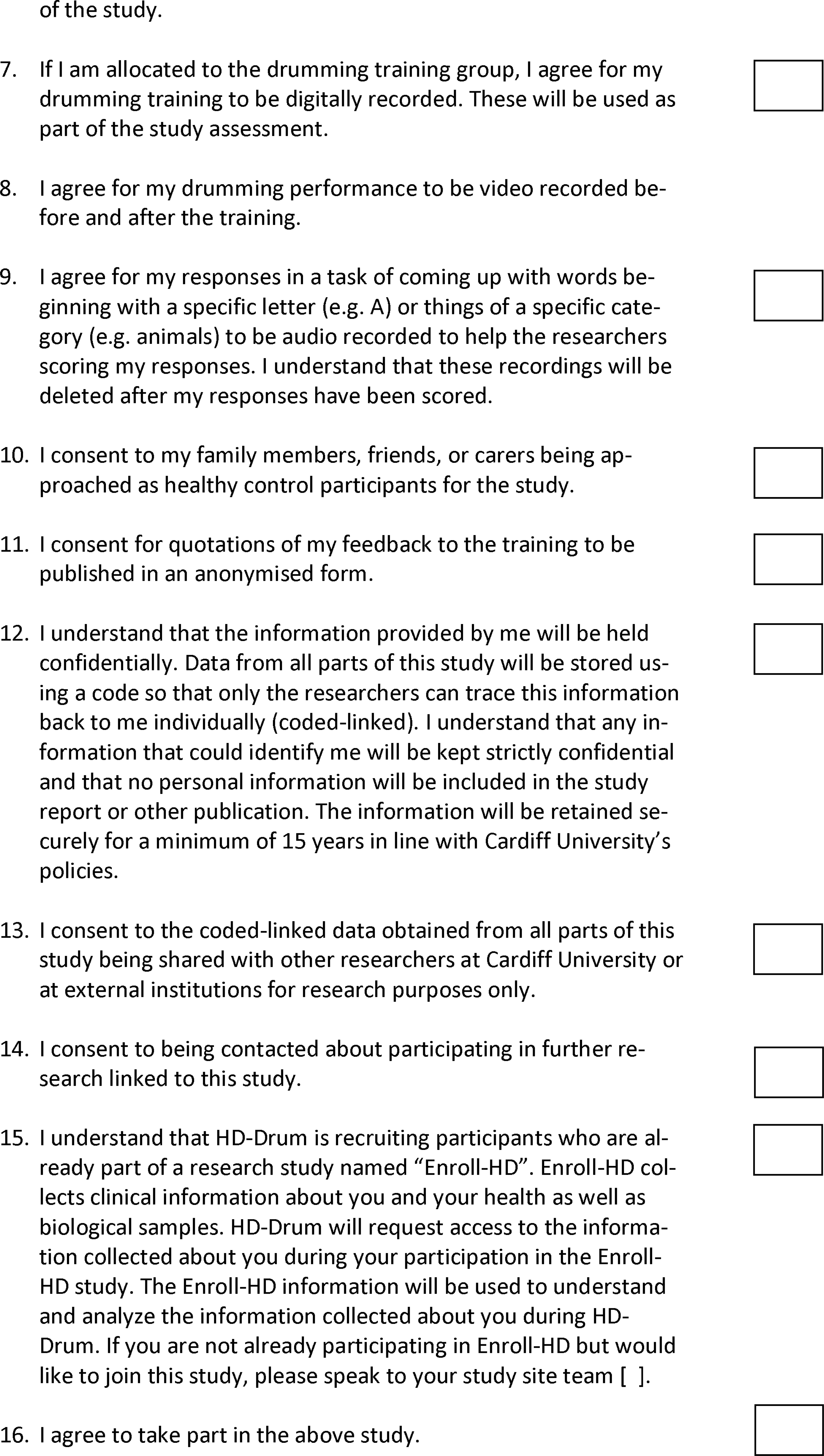
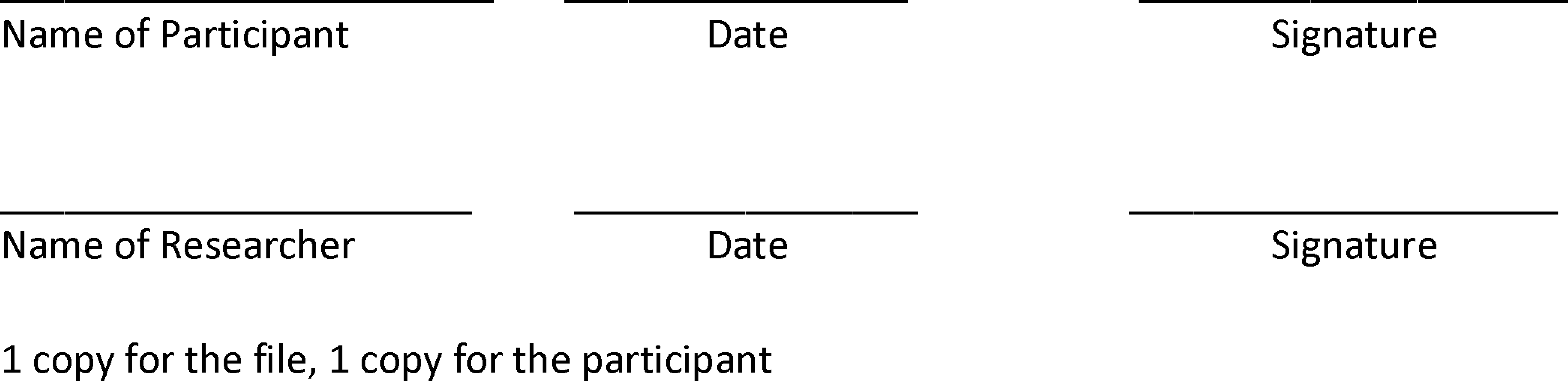

## Notes

### Competing Interest Statement

The authors have declared no competing interest.

### Clinical Trial

ISRCTN 11906973

### Author Declarations

The study has received favourable ethical opinion from the Wales Research Ethics Committee 2 (REC reference: 22/WA/0147) and is sponsored by Cardiff University (SPON1895-22) (Research Integrity, Governance and Ethics Team, Research & Innovation Services, Cardiff University, 2nd Floor, Lakeside Building, University Hospital of Wales, Cardiff, CF14 4XW).

